# HIV Phylogenetics Reveals Overlapping Transmission Networks among Cities and Key Populations in Pakistan

**DOI:** 10.1101/2021.12.14.21267743

**Authors:** Adriano de Bernardi Schneider, François Cholette, Yann Pelcat, Aaron G. Lim, Peter T. Vickerman, Adiba Hassan, Laura H. Thompson, James F. Blanchard, Faran Emmanuel, Tahira Reza, Nosheen Dar, Nadeem Ikram, Richard Pilon, Chris Archibald, Jeffrey B. Joy, Paul Sandstrom, Joel O. Wertheim

## Abstract

The first case of HIV in Pakistan was documented in 1987, with multiple subtypes and circulating recombinant forms being introduced and currently circulating in the country. Since then, there has been a shift in the country from a low prevalence/high risk to a high-risk concentrated epidemic. Pakistan’s epidemic is concentrated among key populations at greater risk of HIV infection including people who inject drugs (PWID), Hijra sex workers (HSW), female sex workers (FSW), male sex workers (MSW), and men who have sex with men (MSM). This study focused on the geographical aspect as well as on the interactions between key populations at higher risk of contracting HIV. We aimed at understanding the behavior of these key populations at a molecular level with high granularity as well as investigating the possibility of multiple HIV-1 introductions in Pakistan. In this cross-sectional biological and behavioral survey, we collected dried blood spots (DBS) for the purposes of seroprevalence estimates and molecular epidemiology from individuals in 17 cities in Pakistan representing four key populations: PWID, HSW, MSW, and FSW. A total of 1153 envelope sequences (reference positions in HXB2: 7860-8274) of HIV were sequenced using a Sanger-based sequencing approach. To identify clusters based on the introduction of the virus in Pakistan from foreign countries we added 3623 publicly available HIV envelope sequences to our dataset. Phylogeographic inference suggests at least 15 independent introductions of the virus into Pakistan, with a total of 12 clusters ranging from 3 to 675 sequences in size containing sequences from Pakistan and neighboring countries exclusively. Our phylogenetic analysis shows a significant degree of connectivity and directionality suggesting broad and overlapping networks of HIV-1 transmission among cities and key populations in Pakistan.

## Introduction

The first case of HIV in Pakistan was documented in 1987 in Karachi (1). Following this first recorded case, multiple subtypes and circulating recombinant forms (CRFs) have been identified and are currently circulating in the country (2–4). Still, before the implementation of integrated biological and behavioral surveillance (IBBS) of key populations in Pakistan in 2005 (5), little was known about the prevalence and associated risk behaviors of HIV in the country (6). Back in 2006, Pakistan was on the brink of a potentially severe epidemic (7), building upon an explosive HIV epidemic among people who inject drugs (PWID) in Karachi dating back as early as 2003 (8). HIV prevalence rapidly increased from 10.8% in 2005 to 38.4% in 2016-17 (5, 9). HIV transmission in large urban centers, such as Karachi and Lahore, has been observed in key populations such as PWID and female sex worker (FSW) (8). In 2011, the reported national HIV prevalence for FSW, Hijra (transgender) sex workers (HSW), male sex workers (MSW), and PWID was 0.8%, 7.2%, 3.1% and 37.8%, respectively (9). Although no significant changes in HIV prevalence was reported during the latest round of IBBS in 2016-17 for PWID (38.4%) and HSW (7.5%), suggesting an established epidemic, the prevalence in FSW and MSW increased approximately two fold from 0.8% to 2.2% and 3.1% to 5.6%, respectively (10). In Pakistan, high risk behaviors such as habitual needle sharing among PWID and frequent condom-less sex among PWID and sex workers, has been associated with an increase in HIV infections in Pakistan (10, 11).

HIV transmission in Asia, although known for its diversity, it is known to start with high rates of infection among PWID followed by sex workers and those who interact with them (7). In Pakistan, a rural to urban exodus as well as with migration of individuals from their native city to other cities in search of work has also been known to be a cause of transmission (7). A 2006 study of migrants observed that the use of condoms when in contact with sex workers was close to 10% (7). A separate study of female sex workers in 2013 found condom usage was <40% (12). Interestingly, although condoms are accessible in Pakistan, using condoms can be challenging given the country’s social conservatism (13).

Scaling up HIV prevention for all key populations in Pakistan is of utmost importance to reduce the overall burden of the disease in the country. A substantial degree of behavioral mixing has led to HIV transmission between key populations, such as PWID and men who have sex with men (MSM), who can then pass on the infection to their spouses and children (14). Along with high transmission rates among PWID, high HIV prevalence among sex workers has been observed, although no direct link of transmission between these populations has been established in Pakistan (15). Recent increases in HIV-1 prevalence among sex workers and MSM may be indicative of overlapping transmission networks among these key populations. It has been well documented that a significant percentage of PWID (>20%) are buying sex from HSW, FSW, and/or MSW (reported condom use very low). Furthermore, a non-negligible proportion of HSW (2.4%), FSW (5.9%), and MSM (4.2%) report injecting drugs (10). Additionally, in other countries in Asia a link between PWID and individuals buying and selling sex has been identified as the center of HIV epidemics (16). Although overlapping injecting and sexual networks have been reported, it hasn’t been shown empirically that this may lead to significant HIV transmission.

In this study, we report the sequencing of 1153 HIV *envelope* sequences from PWID and sex workers across 17 Pakistani cities. We investigate their phylogenetic relationships and transmission dynamics between key populations within Pakistan and ascertain the minimum number of introductions of HIV-1 in the country responsible for these clusters.

## Materials and Methods

### Data collection

Behavioral data and biological samples were collected in 2011 from individuals representing key populations at greater risk of contracting HIV including FSW, HSW, MSW, and PWID across 17 cities in Pakistan (Table 1). The sampling strategy was based on detailed mapping exercises of key populations in Pakistan undertaken prior to surveys and biological specimen collection. The mapping methodology is described in greater detail elsewhere (11). Field teams were dispatched across Pakistan to conduct the actual surveys and biological specimen collection. After obtaining informed consent, participants were interviewed using a structured questionnaire to collect information pertaining to socio-demographics, drug use practices, social networks, sexual behavior, and utilization of services for HIV testing and care. Once the interview was completed, participants were tested for HIV through the use of dried blood spot (DBS) specimens collected by the standard finger prick method using self-retracting lancets. DBS specimen cards were air dried for a minimum of 3 hours at ambient temperature prior to storage at room temperature in sealed gas-impermeable bags containing desiccant pouches. All DBS specimen cards were screened for HIV-1 at the National HIV Reference Laboratory at the National AIDS Programme (Islamabad, Pakistan) and the Armed Institute of Pathology (AFIP; Rawalpindi, Pakistan). All HIV-1 reactive cards and approximately 10% of HIV-1 non-reactive cards (used as quality control) were shipped to the National HIV and Retrovirology Laboratory (NHRL; Winnipeg, Manitoba, Canada) for HIV-1 envelope gene sequencing using Sanger sequencing.

**Table 1.** Sample distribution of collected HIV positive dried blood cards in Pakistan.

| State/Province | City | N | FSW | HSW | MSW | PWID |
| --- | --- | --- | --- | --- | --- | --- |
| Sindh | Dadu | 20 | 0 | 0 | 0 | 20 |
| Punjab | Dera Ghazi Khan | 130 | 2 | 0 | 0 | 128 |
| Punjab | Faisalabad | 155 | 0 | 11 | 0 | 144 |
| Punjab | Gujrat | 62 | 0 | 0 | 0 | 62 |
| Sindh | Karachi | 159 | 2 | 29 | 17 | 111 |
| Punjab | Lahore | 110 | 2 | 11 | 6 | 91 |
| Sindh | Larkana | 96 | 2 | 37 | 9 | 48 |
| Punjab | Multan | 82 | 1 | 4 | 7 | 70 |
| Punjab | Pakpattan | 5 | 0 | 0 | 0 | 5 |
| Khyber Pakhtunkhwa | Peshawar | 35 | 0 | 4 | 0 | 31 |
| Balochistan | Quetta | 27 | 0 | 5 | 3 | 19 |
| Punjab | Rahimyar Khan | 28 | 0 | 0 | 0 | 28 |
| Punjab | Rawalpindi | 14 | 0 | 12 | 2 | 0 |
| Punjab | Sargodha | 134 | 0 | 13 | 0 | 121 |
| Sindh | Sukkur | 69 | 1 | 22 | 0 | 46 |
| Balochistan | Turbat | 27 | 0 | 0 | 0 | 27 |

N, total number of sequences; FSW, Female sex worker; HSW, Hijra sex worker; MSW, male sex worker; PWID, Person who inject drugs

A routine, in-house HIV-1 genotyping assay was used to sequence a portion of the *envelope* gene (HXB2 reference positions: 7860-8274). Total nucleic acid was isolated from a single DBS (approximately 75 *µ*L of whole blood) using an automated, magnetic silica-based, NucliSENS easyMAG (bioMerieux) according to the manufacturer’s specific protocol B. Each DBS was lysed for 1 hour at room temperature with gentle agitation in 2 mL of NucliSENS lysis buffer. Nucleic acid was eluted in 50 *µ*L of NucliSENS extraction buffer 3 and stored at −80C until further testing. The envelope gene was amplified by reverse transcriptase PCR (RT-PCR) followed by nested PCR. RT-PCR thermal cycling conditions consisted of 50C/10 min, 98C/2 min followed by 40 cycles of 98C/10 sec, 50C/10 sec, 72C/60 sec and a final extension of 72/5 min. Each reaction contained 10.5 *µ*L of water, 25 *µ*L of SuperScript 2X Reaction Mix, 2 *µ*L of primers gp41F1 and gp41R1 (200 nM final concentration), 0.5 *µ*L of SuperScript IV RT mix, and 10 *µ*L of template. Nested PCR thermal cycling conditions consisted of 94C/2 min followed by 40 cycles of 94C/15 sec, 60C/15 sec, 68C/30 sec and a final extension of 68/5 min. Each nested PCR reaction contained 16 *µ*L of water, 25 *µ*L of 2X Platinum II Master Mix, 2 *µ*L of primer gp46F2 and gp48R2 (400 nM final concentration), and 5 *µ*L of template. Primer sequences for gp41 were the following: gp41F1:CTTAGGAGCAGCAGGAAGCACTATGGG; gp41R1:AACGACAAAGGTGAGTATCCCTGCCTAA; gp41F2:ACAATTATTGTCTGGTATAGTGCAACAGCA; gp41R2:TCCTACTATCATTATGAATATTTTTATATA. Sanger sequencing was performed using BigDye Terminator v3.1 cycle sequencing kit with nested PCR primers. Sequence analysis and contig assembly were performed using Geneious software.

A total of 1153 HIV-1 partial *envelope* sequences were sequenced using the Sanger-based sequencing approach described above in Winnipeg at the JC Wilt Infectious Diseases Research Centre, National HIV and Retrovirology Laboratories, Public Health Agency of Canada. The sequences were deposited on GenBank (Accession numbers: MW304006MW305158).

Ethical approval was obtained from the Hope International and Health Canada/Public Health Agency of Canada Ethical Review Board (REB-2010-0044). Informed consent was obtained verbally from study participants for the behavioral survey and biological sampling components of this study. Verbal consent was chosen due to the varying levels of literacy among study participants as well as to protect their identities. Each consent was documented by a member of the data collection team.

### Maps of prevalence

Cities with prevalence data greater than 0 were visualized as heatmaps. To create the heatmaps, data points were projected using ArcMap 10.7. A Jenk’s natural break method was used to classify the raster into five categories for cartographic representation.

### Molecular data and sequence analyses

Multiple sequence alignment was performed using MAFFT v7.215 (17) under default settings. The alignments were visualized in AliView and ragged edges were trimmed manually when necessary (18). We classified the HIV sequences into subtypes using a combination of COMET (19) and phylogenetic inference using IQ-TREE (20). For this phylogenetic analysis we included 170 reference *envelope* sequences obtained from the Los Alamos National Laboratories HIV databases (LANL https://www.hiv.lanl.gov) encompassing all available subtypes and CRFs. Results from COMET analysis were compared to the clades to which each sequence clustered with the reference sequence.

We downloaded 3623 HIV *envelope* sequences from the LANL HIV sequence database, encompassing all the available A1, G and CRFs 02,35 and 63 from the HIV database at the time of search, creating a large diverse dataset with sequences from around the world from the same subtypes identified as the most prevalent on our dataset to identify individual clusters of HIV introduction in Pakistan (see supplementary data). All the geographic metadata available in the public domain for these envelope sequences was compiled (see supplementary data). We separated the Pakistani sequences from the initial dataset into twelve subset clusters to study the relationships between the taxon within each individual cluster. These subsets encompassed all sequences belonging to Pakistan and neighboring Asian countries.

### Phylogenetic tree search

To determine the HIV clusters we performed a maximum-likelihood tree search method using FastTree (21) under the GTRCAT model of nucleotide substitution with 100 bootstraps and the -pseudo, -spr and mlacc options for improved accuracy. Clades containing only Pakistani sequences with a minimum of 85% bootstrap value were selected as individual clusters. When available, neighboring Asian countries that were under the same bootstrap threshold were also included as part of the clusters. The final phylogenetic tree was rendered using Evolview v2 (22).

### Genetic distance

To calculate the genetic distance within each individual Pakistani clade we estimated the evolutionary divergence over sequence pairs within groups as implemented in MEGA X under default settings (23, 24). The analyses were conducted using the Maximum Composite Likelihood model (25).

### Phylogeography of individual clusters

BEAST v.10.4 was used to attain the molecular dating and phylogeographic diffusion of the individual clusters (26). The phylogeographic models were used to estimate both the transmission between locations and key populations. We performed the phylogeographic diffusion in discrete space analysis using BSSVS (a Bayesian Stochastic Search Variable Selection) (27). We calculated the substitution model for each cluster using ModelFinder as implemented on IQ-TREE (20), and used the Skyride coalescent tree prior (28). Results were visualized in Tracer (29) and convergence was determined by Effective Sample Sizes of at least 200 per statistic. Strict clock and uncorrelated relaxed lognormal clock (URL) were tested for each cluster. We ran the model in triplicate with a minimum Markov chain Monte Carlo (MCMC) length of 100 million for each cluster and 10% burn-in to each original MCMC. The final maximum clade credibility trees with metadata annotation were rendered using FigTree (30).

Additionally, Bayes Factor (BF) and Markov Jumps were calculated for each individual cluster. We wrote a script to extract both Bayes Factor and Markov Jumps from BEAST log files (available at https://github.com/abschneider/phylogeography-density-plots). We calculated the BF for each individual cluster for the transition between cities and key populations. Results were compiled and a table summarizing all transmissions between cities and key populations was created. We used a BF threshold of *≥* 5 to accept a transmission as significant (31).

### Phylogeography visualization

Individual networks were visualized using the MCC trees on StrainHub v1.0.1 (32). The combined transmission table was used to generate a network map for the transmission between cities. The network map was created using ESRI ArcMap version 10.8. Country administrative boundaries GADM version 3.6 (https://gadm.org/) were used as a basemap.

### Data availability

All supplementary data can be found at GitHub (https://github.com/abschneider/Paper_HIV_Pakistan).

The Bayes Factor script is available at GitHub (https://github.com/abschneider/phylogeography-density-plots).

## Results

### Subtype classification

Subtype classification analysis using COMET (19) and phylogenetic inference using IQTREE (33) indicates at least six major subtypes/CRFs (Circulating Recombinant Forms) represented on our dataset: A1, G, C, D, CRF02_AG and CRF35. The sequences that had mismatches between both COMET and the phylogenetic analysis subtyping analyses were labeled as “unknown”. When results from COMET suggested the possibility of recombinants, and these were confirmed at the phylogenetic analysis, we classified according to the phylogenetic analysis result (i.e., “A1 (check for 35_AD)” / “35_AD” = 35_AD)(see Supplementary Material). Higher prevalence subtypes were identified on the dataset as A1 (64%), G (5%), CRF02_AG (2%) and CRF35 (4%), 24% of the dataset sequences were identified as unknown.

### Nucleotide divergence

The nucleotide divergence found within each individual clade was relatively low (< 0.06) with an average of 0.04, with the exception of clade IV (0.12) (Figure 2). Clade IV (N=18), the clade with highest nucleotide divergence, is the only one with sequences identified as belonging to subtype C, which presents low prevalence over the entire dataset (>1%).

**Fig. 1.**
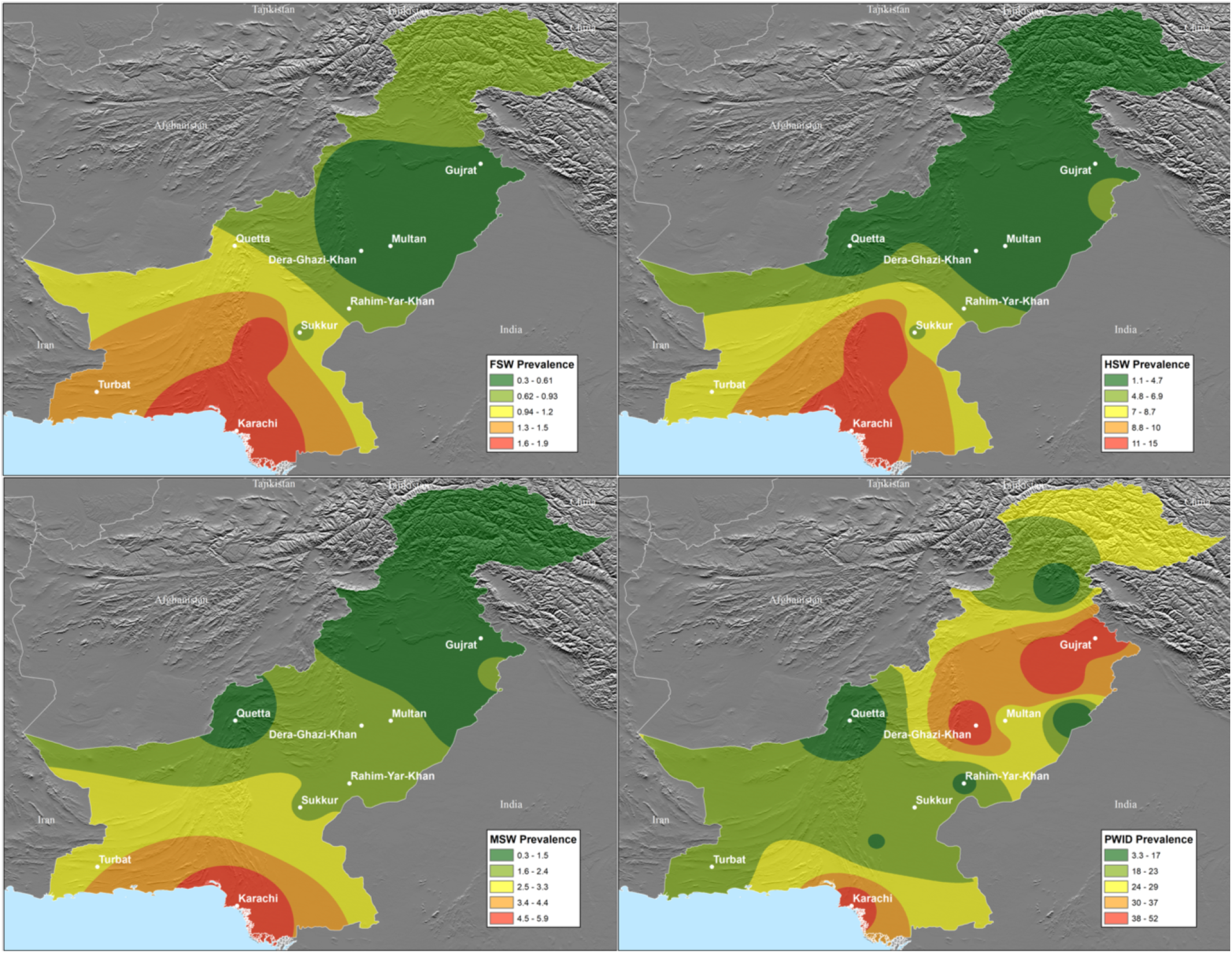
Location of the study population and prevalence of HIV within the area in distinct key populations (9). Top left: Female Sex Workers (FSW); Top Right: Hijra Sex Worker; Bottom Left: Male Sex Worker (MSW); Bottom Right: People Who Inject Drugs (PWID).

**Fig. 2.**
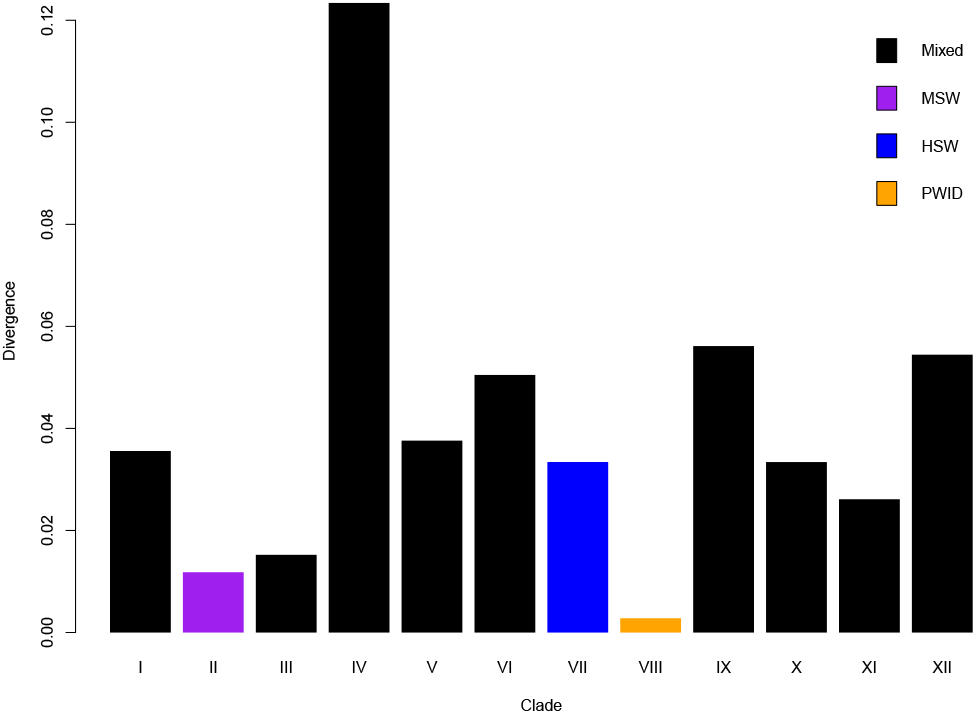
Estimates of average evolutionary divergence over sequence pairs within groups. The number of base substitutions per site from averaging over all sequence pairs within each group are shown. Analyses were conducted using the Maximum Composite Likelihood model (25). This analysis involved 1307 nucleotide sequences.

### Phylogenetic analysis suggests multiple introductions of HIV in Pakistan

The maximum likelihood tree has 12 clades containing Pakistani sequences and three single terminal sequences from Pakistan (Figure 3). These independent clades include LANL sequences from countries around the world which separate them and indicate multiple independent introductions of HIV into Pakistan (see Supplementary Material for complete tree). The sequences clustered into a major cluster (I) with 675 sequences, six medium size clusters with 38-226 sequences (III, V, VI, IX, XI and XII) and five minor clusters with 3-18 sequences (II, IV, VII, VIII and X)(Table 2). Cluster XII contains sequences from Iran and Afghanistan, indicating transmission events happening between individuals across the border countries. The three sequences that did not cluster with other Pakistani sequences were ignored for individual cluster/transmission network analysis.

**Fig. 3.**
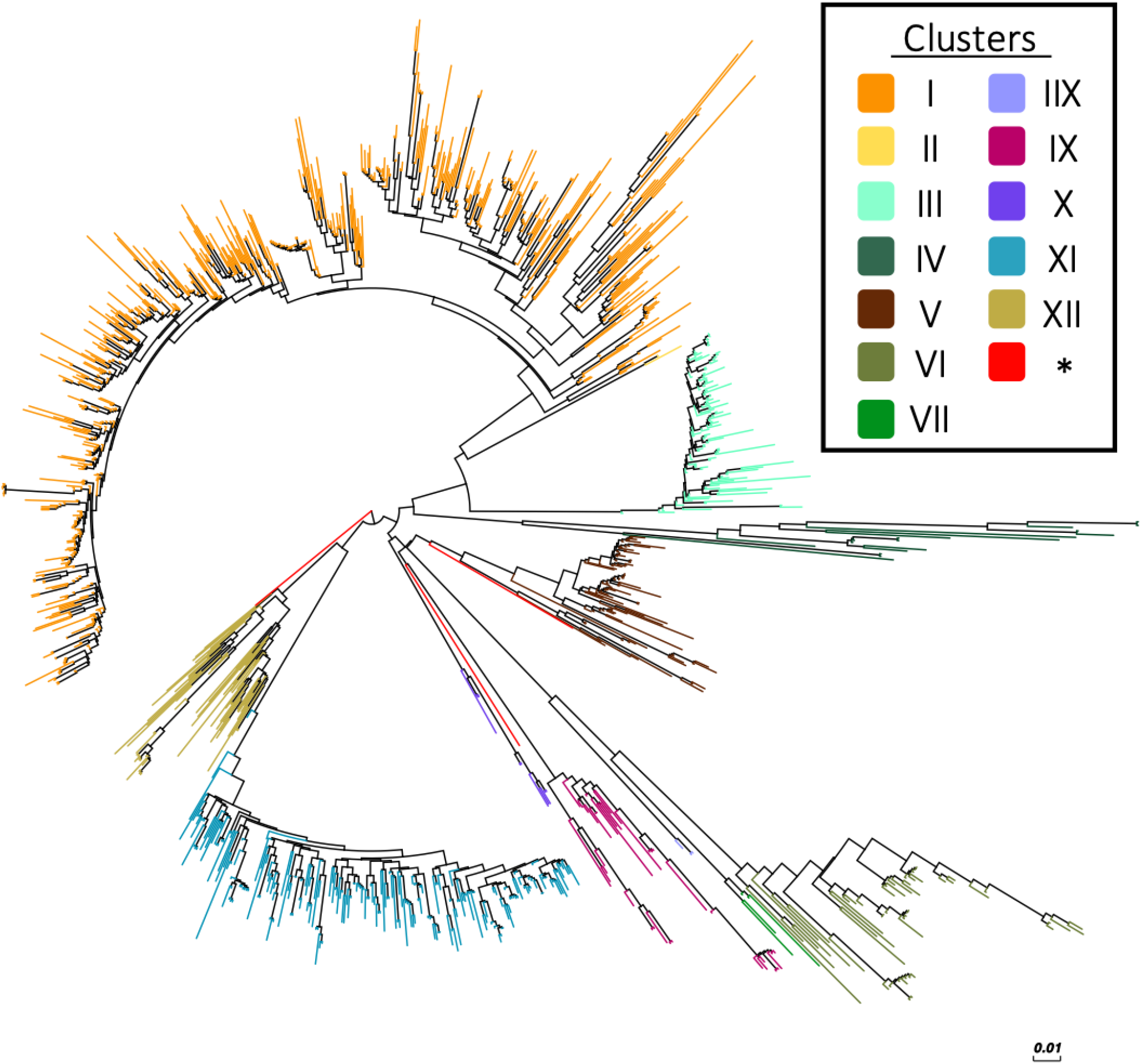
Maximum-likelihood tree of HIV envelope genes representing independently introduced HIV strains in Pakistan. Subtree is shown, LANL sequence terminals were omitted to aid the visualization, complete tree can be seen on supplementary material.

**Table 2.**
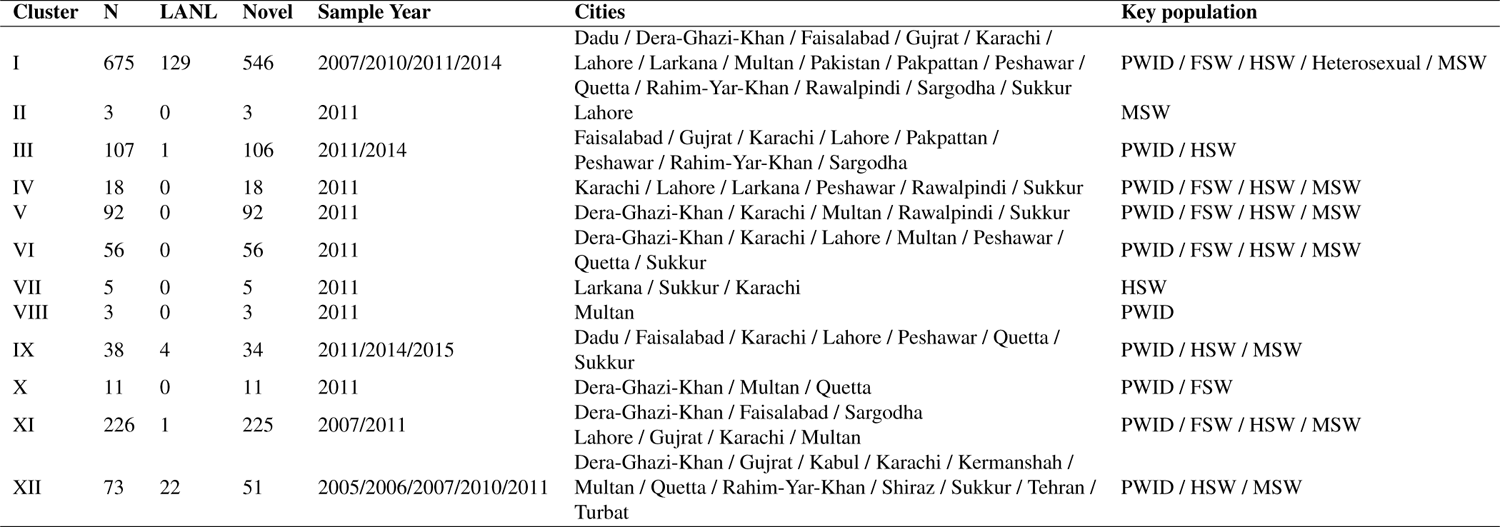
Cluster distribution of samples

| Cluster | N | LANL | Novel | Sample Year | Cities | Key population |
| --- | --- | --- | --- | --- | --- | --- |
| I | 675 | 129 | 546 | 2007/2010/2011/2014 | Dadu / Dera-Ghazi-Khan / Faisalabad / Gujrat / Karachi / Lahore / Larkana / Multan / Pakistan / Pakpattan / Peshawar / Quetta / Rahim-Yar-Khan / Rawalpindi / Sargodha / Sukkur | PWID / FSW / HSW / Heterosexual / MSW |
| II | 3 | 0 | 3 | 2011 | Lahore | MSW |
| III | 107 | 1 | 106 | 2011/2014 | Faisalabad / Gujrat / Karachi / Lahore / Pakpattan / Peshawar / Rahim-Yar-Khan / Sargodha | PWID / HSW |
| IV | 18 | 0 | 18 | 2011 | Karachi / Lahore / Larkana / Peshawar / Rawalpindi / Sukkur | PWID / FSW / HSW / MSW |
| V | 92 | 0 | 92 | 2011 | Dera-Ghazi-Khan / Karachi / Multan / Rawalpindi / Sukkur | PWID / FSW / HSW / MSW |
| VI | 56 | 0 | 56 | 2011 | Dera-Ghazi-Khan / Karachi / Lahore / Multan / Peshawar / Quetta / Sukkur | PWID / FSW / HSW / MSW |
| VII | 5 | 0 | 5 | 2011 | Larkana / Sukkur / Karachi | HSW |
| VIII | 3 | 0 | 3 | 2011 | Multan | PWID |
| IX | 38 | 4 | 34 | 2011/2014/2015 | Dadu / Faisalabad / Karachi / Lahore / Peshawar / Quetta / Sukkur | PWID / HSW / MSW |
| X | 11 | 0 | 11 | 2011 | Dera-Ghazi-Khan / Multan / Quetta | PWID / FSW |
| XI | 226 | 1 | 225 | 2007/2011 | Dera-Ghazi-Khan / Faisalabad / Sargodha<br>Lahore / Gujrat / Karachi / Multan | PWID / FSW / HSW / MSW |
| XII | 73 | 22 | 51 | 2005/2006/2007/2010/2011 | Dera-Ghazi-Khan / Gujrat / Kabul / Karachi / Kermanshah / Multan / Quetta / Rahim-Yar-Khan / Shiraz / Sukkur / Tehran / Turbat | PWID / HSW / MSW |

### Bayesian inference and transmission dynamics of HIV clusters

Given the lack of temporal information for most of the clusters, we selected the cluster with most variable temporal data, cluster XII (Table 2), to calculate the clock and use it as the clock prior for all other individual cluster runs.

We performed analysis on ten of the twelve clusters. Clusters II and VIII only contained a single location and a single key population, thus, they were omitted from this analysis. The results of the individual analysis can be visualized in the supplementary material.

The combined data of each individual cluster shows transmission pattern between cities show three major cities for transmission given our current data, Karachi, Multan and Lahore, with Karachi mostly acting like a sink (over 70% of inbound transmission), Multan as a hub (inbound transmission equal to outbound transmission) and Lahore as a source (over 70% of outbound transmission) for HIV transmission (Figure 5). The transmission between key populations indicates connectivity between PWID and sex workers, which was observed in multiple clusters (I, III, IX, X and XI) as well as between FSW, MSW and HSW (I,IV,V,VI,IX) with Bayes Factor *≥* 5 (Figure 4).

**Fig. 4.**
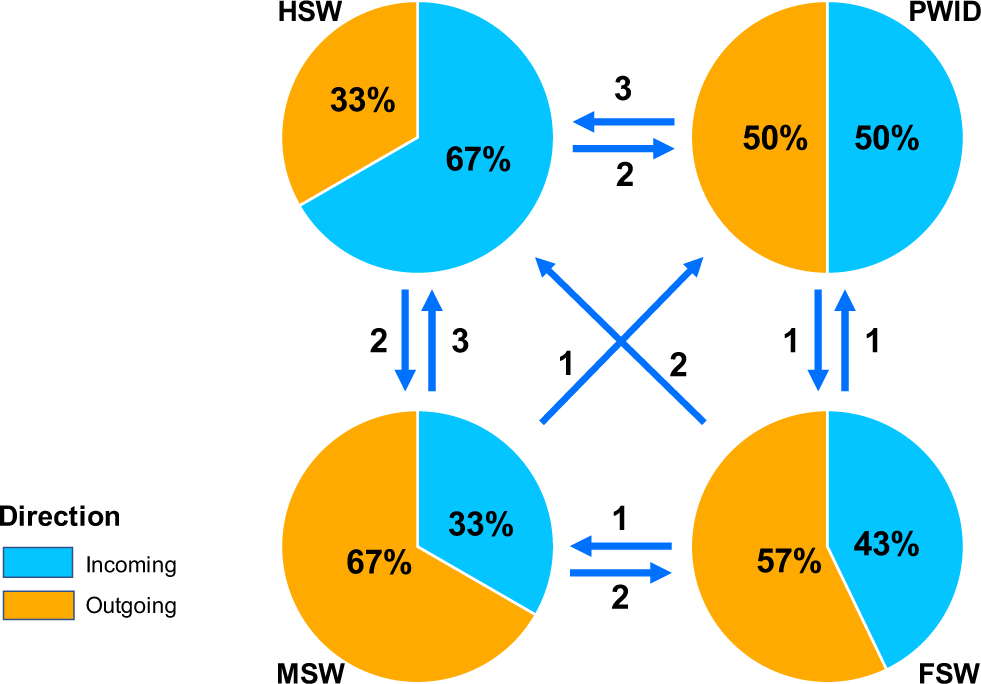
Summarized HIV-1 transmission network between key populations in Pakistan. Arrows represent the direction of the transmission and numbers represent the number of observed transmissions between key populations that were supported by a Bayes Factor threshold ≥ 5.

**Fig. 5.**
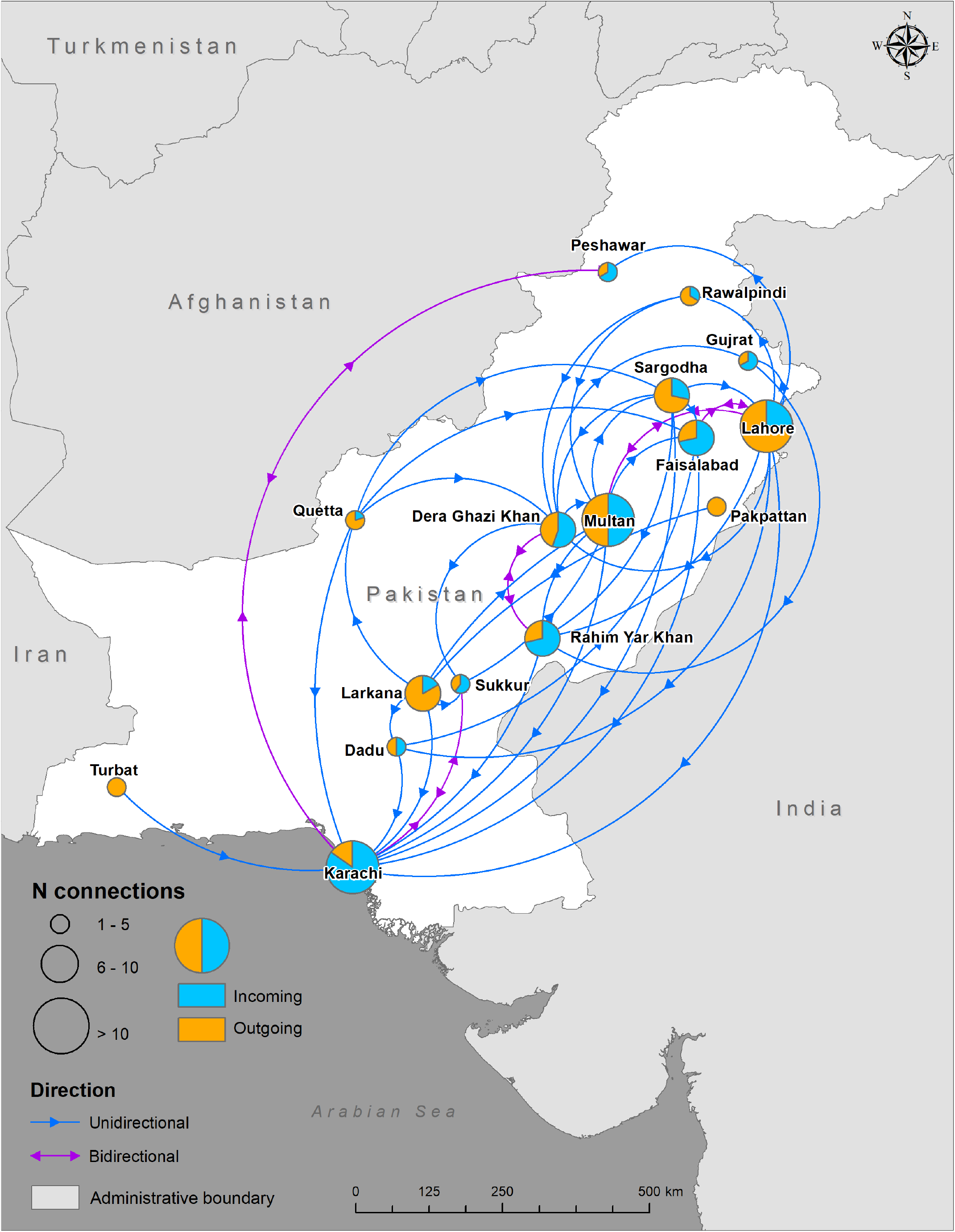
Summarized HIV-1 transmission network in Pakistan. Pie charts size represent number of connections between cities and colors represent incoming and outgoing transmissions between cities. Transmission network was generated using Bayes Factor threshold *≥* 5. Network map was created using ESRI ArcMap version 10.8. Country administrative boundaries GADM version 3.6 () were used as a basemap.

Due to each individual cluster represent only the individual spread of that particular introduction in Pakistan, we elected to combine all the results in order to visualize the dynamics of transmission among all clusters and have a picture of the overall transmission pattern of HIV in Pakistan with our dataset. The rationale behind this is that regardless of separate introductions in the country, the dynamics of HIV occurs between hosts, rather than the virus, thus, combining the clusters give a better picture of the transmission pattern happening between the key populations in this study rather than each cluster separately.

## Discussion

We characterized multiple introductions of HIV-1 into Pakistan, representing numerous subtypes: A1, G, C, D, CRF02_AG, and CRF35_AD. Our subtyping results are aligned with previous research which suggests that CRF02_AG/A1 recombinants predominate the prototype CRF02_AG viruses, while subtype A1 still dominates the virus population in Pakistan (2). According to Chen et al. (2), the majority of subtype A viruses in Pakistan were the result of a single introduction. In contrast, our results indicate the presence of subtype A1 in multiple independent clusters, proving that the former conclusion was hampered by the lack of evidence and highlighting the importance of a thorough phylogenetic analysis of HIV-1 genomic data in Pakistan. The presence of at least twelve introductions into the country of multiple subtypes, observed with the current dataset, validates the importance of cross border transmission in the onset of infectious diseases epidemics (34). The larger nucleotide divergence present at cluster IV, which may be explained given the low number of sequences for the subtypes present in our dataset for that clade (B, C and D), give initial evidence to support an even higher number of introductions to the country, although further studies with focus on these subtypes should be performed to allow a better understanding of their introductions into Pakistan.

Prevalence of HIV in Pakistan is concentrated in vulnerable populations such as PWID and sex workers, whereas prevalence of HIV in the general population seems to be underreported (35). As observed in our results, HIV transmission is not occurring in silos, with PWID and sex workers being interconnected as well as multiple cities across the whole country (Figures 4 and 5). Geographically, Lahore and Karachi are seen in our analysis as the main cities in Pakistan responsible for the spread of HIV-1 among these key populations. These two are major cities, with Karachi being the economic center of Pakistan, and Lahore being considered the cultural center with a significant sex worker population. A previous study pointed Larkana, the 15th most populous city in Pakistan, as the center of the HIV epidemic among PWID, indicating that the dynamics of the disease seems to change when additional key populations are included in the analysis (36). Nevertheless, to date, no study has connected these two key populations (sex workers and PWID) at a molecular level. Previous studies restricted to epidemiological field studies have suggested considerable overlapping HIV transmission occurring between PWID and sex workers and that overlapping transmission between these key populations existed based on surveys (15).

Neighboring countries, such as Iran, have reported HIV transmission by truck drivers (37). Commercial truck drivers in routes connecting main cities in Pakistan are also known to have contact with sex workers and potentially becoming an additional key population prompting long distance geographic spread of HIV (8, 38). Most HIV prevention efforts in Pakistan have focused on PWID and for several years this was not seem as of major concern given that HIV prevalence appeared to be relatively stable among sex workers (5, 39– 41). Recently, the increase of HIV prevalence observed among sex workers could also be explained by lack of interventions in these key populations as well as an overlapping of sexual/injecting networks with PWID (high prevalence). Although Pakistan has made significant advancements towards its HIV/AIDS response, political and social challenges enable the continuous transmission of HIV between these populations (42).

By combining the observed behavior of each individual introduction, we were able to understand the overall pattern of HIV-1 transmission among the key populations of this study. Given that individuals are unaware to which HIV subtype they are at risk of contracting, individual cluster dynamics are irrelevant from the perspective of understanding transmission networks in general when not trying to trace individual contact networks. This study focused on the geographical aspect as well as on the interactions between key populations at higher risk of contracting HIV.

## Conclusion

Our findings document at least 15 distinct introductions of HIV into Pakistan, with a total of 12 clusters representing onward transmission within Pakistan. The combined information from the phylogeographic analysis of the individual clusters used to estimate the transmission between key populations suggests constant transmission of HIV between PWID and sex workers. Closely related sequences from Afghanistan and Iran, suggests repeated movement of HIV across these international borders, which could be explained by drug trade and refugees cross between the countries. Nevertheless, our results uncover only the surface of the epidemic given the limitation of what could be explored with our current dataset. Further studies with fine grained data should perform to understand better local transmission networks to allow for more comprehensive public health interventions and surveillance among key populations in Pakistan.

## Data Availability

All supplementary data can be found at GitHub (https://github.com/abschneider/Paper_HIV_Pakistan). The Bayes Factor script is available at GitHub (https://github.com/abschneider/phylogeography-density-plots).

## Acknowledgements

We thank the surveillance team for their efforts in mapping, data collection, specimen collection, and data entry. We also thank the participants who took part in the surveys for their time and contribution.

